# Development and Validation of a Deep Learning Model for Detecting Signs of Tuberculosis on Chest Radiographs among US-bound Immigrants and Refugees

**DOI:** 10.1101/2024.02.27.24303429

**Authors:** Scott Lee, Shannon Fox, Raheem Smith, Kimberly A. Skrobarcek, Harold Keyserling, Christina R. Phares, Deborah Lee, Drew L. Posey

## Abstract

Immigrants and refugees seeking admission to the United States must first undergo an overseas medical exam, overseen by the US Centers for Disease Control and Prevention (CDC), during which all persons ≥15 years old receive a chest x-ray to look for signs of tuberculosis. Although individual screening sites often implement quality control (QC) programs to ensure radiographs are interpreted correctly, the CDC does not currently have a method for conducting similar QC reviews at scale.

We obtained digitized chest radiographs collected as part of the overseas immigration medical exam. Using radiographs from applicants 15 years old and older, we trained deep learning models to perform three tasks: identifying abnormal radiographs; identifying abnormal radiographs suggestive of tuberculosis; and identifying the specific findings (e.g., cavities or infiltrates) in abnormal radiographs. We then evaluated the models on both internal and external testing datasets, focusing on two classes of performance metrics: individual-level metrics, like sensitivity and specificity, and sample-level metrics, like accuracy in predicting the prevalence of abnormal radiographs.

A total of 152,012 images (one image per applicant; mean applicant age 39 years) were used for model training. On our internal test dataset, our models performed well both in identifying abnormalities suggestive of TB (area under the curve [AUC] of 0.97; 95% confidence interval [CI]: 0.95, 0.98) and in estimating sample-level counts of the same (−2% absolute percentage error; 95% CIC: −8%, 6%). On the external test datasets, our models performed similarly well in identifying both generic abnormalities (AUCs ranging from 0.89 to 0.92) and those suggestive of TB (AUCs from 0.94 to 0.99). This performance was consistent across metrics, including those based on thresholded class predictions, like sensitivity, specificity, and F1 score.

Strong performance relative to high-quality radiological reference standards across a variety of datasets suggests our models may make reliable tools for supporting chest radiography QC activities at CDC.

## Introduction

Tuberculosis is an infectious disease caused by *Mycobacterium tuberculosis* (MTB) that typically affects the lungs.^11^Those who are infected but do not show symptoms have latent tuberculosis infection (LTBI) and may never develop tuberculosis disease. LTBI is not infectious but still needs to be treated to prevent the progression into tuberculosis disease. Tuberculosis disease causes coughing, chest pain, fatigue, weight loss, fever, and many other symptoms, and is contagious.^22^ It is the 13^th^ leading cause of death in the world, and the second leading infectious killer after COVID-19.^1^ In the United States, tuberculosis rates have been declining, and the tuberculosis incidence rate for 2021 was 2.4 cases per 100,000 persons, with the majority of reported cases occurring among non-US–born persons (71.4%). Non-US born persons had an incidence rate 15.8 times higher (12.5 cases per 100,000) when compared to US-born persons (0.8 cases per 100,000).^3^

Every year, approximately 550,000 immigrants and refugees apply to enter the United States. The Division of Global Migration Health (DGMH) within the Centers for Disease Control and Prevention (CDC) has regulatory responsibility to oversee the medical examinations of these applicants. The examinations are conducted overseas in accordance with CDC DGMH’s Technical Instructions for panel physicians. All panel physicians are licensed local medical doctors on an agreement with the US Department of State to perform these examinations, and many are affiliated with the International Organization for Migration (IOM), an intergovernmental agency under the United Nations system that supports migrants. IOM works closely with US Department of State and CDC to ensure the healthy migration of US-bound immigrants and refugees.

DGMH’s Technical Instructions for tuberculosis seek to prevent disease importation by detecting and treating infectious tuberculosis before arrival, and to reduce tuberculosis-related morbidity and mortality in these populations. Requirements include a medical history and physical examination. All applicants 15 years and older receive chest x-rays, and anyone with a chest x-ray suggestive of tuberculosis, signs or symptoms suggestive of tuberculosis, or known HIV, then has three sputum specimens collected for smears and cultures.^4,5^

In September 2018, DGMH began receiving digital copies of chest x-ray images from panel sites. This was due to the rollout of the eMedical system, an electronic health processing system that collects data from the required overseas immigrant examinations. In 2018 alone, 124,551 images for 521,270 applicants were collected, raising the possibility of using machine learning methods to complement DGMH’s already effective oversight for the radiologic components of tuberculosis screening for US-bound immigrants and refugees.^6^

### Brief overview of AI for Chest radiography

Artificial intelligence (AI), especially as enabled by deep learning algorithms, has been widely studied for applications in medical imaging. Examples include diabetic retinopathy^7^, cardiovascular risk prediction^8^, cancer histopathology^9–11^, and imaging for musculoskeletal^12,13^, cardiac^14^, and pulmonary^15^conditions. Models are typically designed for diagnostic tasks, like segmenting anatomical structures or indicating the presence of disease, but they have also been designed for prognostic tasks, like predicting survival time for patients from histopathology whole-slide images.^16^

In chest imaging, applications have generally focused on identifying abnormalities associated with specific diseases, like pneumonia,^17,18^, COVID-19^19^, lung cancer^20,21^, and tuberculosis.^15,17,22^ Recent work^23,24^ has broadened the scope to include abnormalities in general. Studies focusing on tuberculosis have ranged from the narrow evaluation of specific models (typically commercial) on relatively small test sets^25,26^ to the development of original algorithms from custom largescale training sets.^23,27,28^ The references standards for these studies are often mixed, comprising radiological findings, clinical diagnoses, microbiological testing, and nucleic acid amplification testing (NAAT).

Of special note, when laboratory tests are used as reference standards, model performance tends to drop relative to performance against a radiological standard; however, a small number of models have met the World Health Organization’s (WHO) Target Product Profile (TPP) for tuberculosis triage tests at 90% sensitivity and at least 70% specificity^26,29^ relative to NAAT or culture, even when testing does not rely on initial radiographic interpretation to identify images with abnormalities (see Khan 2020 and Qin 2021, where all study participants received both a chest x-ray and either a GeneXpert MTB/RIF test or a sputum culture upon enrollment).^26,30^

### Project goal

The primary use-cases of models in the literature have mostly been clinical decision support and workflow improvement, with special emphasis on individual-level classification performance (often as measured by AUC), interpretability, and usability. With respect to TB, emphasis has also been placed on the potential benefit for models to bolster TB screening and diagnosis in low-resource settings, e.g., by rank-ordering radiographs in batches by their probability of disease to guide manual review. For this project, we evaluated our models’ ability to meet these goals, and we also sought to evaluate their performance in estimating sample-level prevalence, i.e., in predicting the number of abnormal x-rays in a given batch. These measures mirror two important operational use-cases of the model in the overseas screening program: supporting panel physicians in providing high-quality initial reads during the exams (an unplanned but potentially impactful application), and enabling DGMH to conduct quality control (QC) with the radiographs once they have been collected (the primary focus of our current project).

To achieve our goals, we trained and validated models for performing three tasks: classifying images as abnormal (Task 1), classifying images as abnormal and suggestive of tuberculosis (Task 2), and identifying the specific abnormalities in the images (Task 3) (we use the same numbering scheme to identify the corresponding models). To meet the two use-cases above, we tested our models on a variety of data sets, both internal and external, and we measured their performance using two operating points, one chosen to optimize individual-level classification performance, and one chosen to optimize accuracy in predicting prevalence. Although we did not formally test abnormality localization methods, e.g., via object detection models, we implemented a number of common saliency methods for visualizing suspected abnormalities on the input images to improve model interpretability and pilot interactive methods for manual review.

## Methods

### Internal dataset curation and description

For our internal datasets (hereafter HaMLET, from our project title, Harnessing Machine Learning to Eliminate Tuberculosis), we obtained an initial convenience sample of 327,650 digitized radiographs from four sources: eMedical, the US Department of State’s immigrant health data system, a web-based application for recording and transmitting immigrant medical cases between the Panel Physicians, US Department of State, and the CDC^31^; the Migrant Management Operational System Application (MiMOSA), the International Organization for Migration’s (IOM) refugee health data system; IOM’s Global Teleradiology and Quality Control Centre (GTQCC); and a small number of individual US immigrant panel sites that screen a relatively high number of applicants with tuberculosis each year (site names are provided in the Acknowledgments). Importantly, all these sites have experienced radiologists and most conduct either double or triple readings on all chest x-ray images as a measure of quality control. Regardless of source, all radiographs were stored as Digital Imaging and Communications in Medicine (DICOM) files, and all radiographic findings were extracted directly from the structured entries in the DS-3030 Tuberculosis Worksheet^32^ instead of from free-text radiology reports by way of natural language processing (NLP).

The set assembled for this project was taken from screenings conducted during a ten-year period from October 2011 to October 2021 and not exclusively from the digitized radiographs routinely received by DGMH since 2018 (Supplemental Table 2 shows the distribution of exams by region and year). The digitized radiographs began in 2018 due to the eMedical rollout, but we also received screenings directly from private immigrant panel sites and from IOM that predated the eMedical rollout. We excluded radiographs from applicants less than 15 years of age (n=52,523), as well as those stored in DICOM files whose pixel arrays were missing, corrupt, or otherwise unreadable by the software we used for extraction (n=107,115) (Figure 1 shows a flow diagram providing a detailed numerical accounting of these two exclusion steps). The remaining 168,012 radiographs constituted our final dataset, which we split into training, validation, and testing portions following the procedure described below.

**Figure 1.**
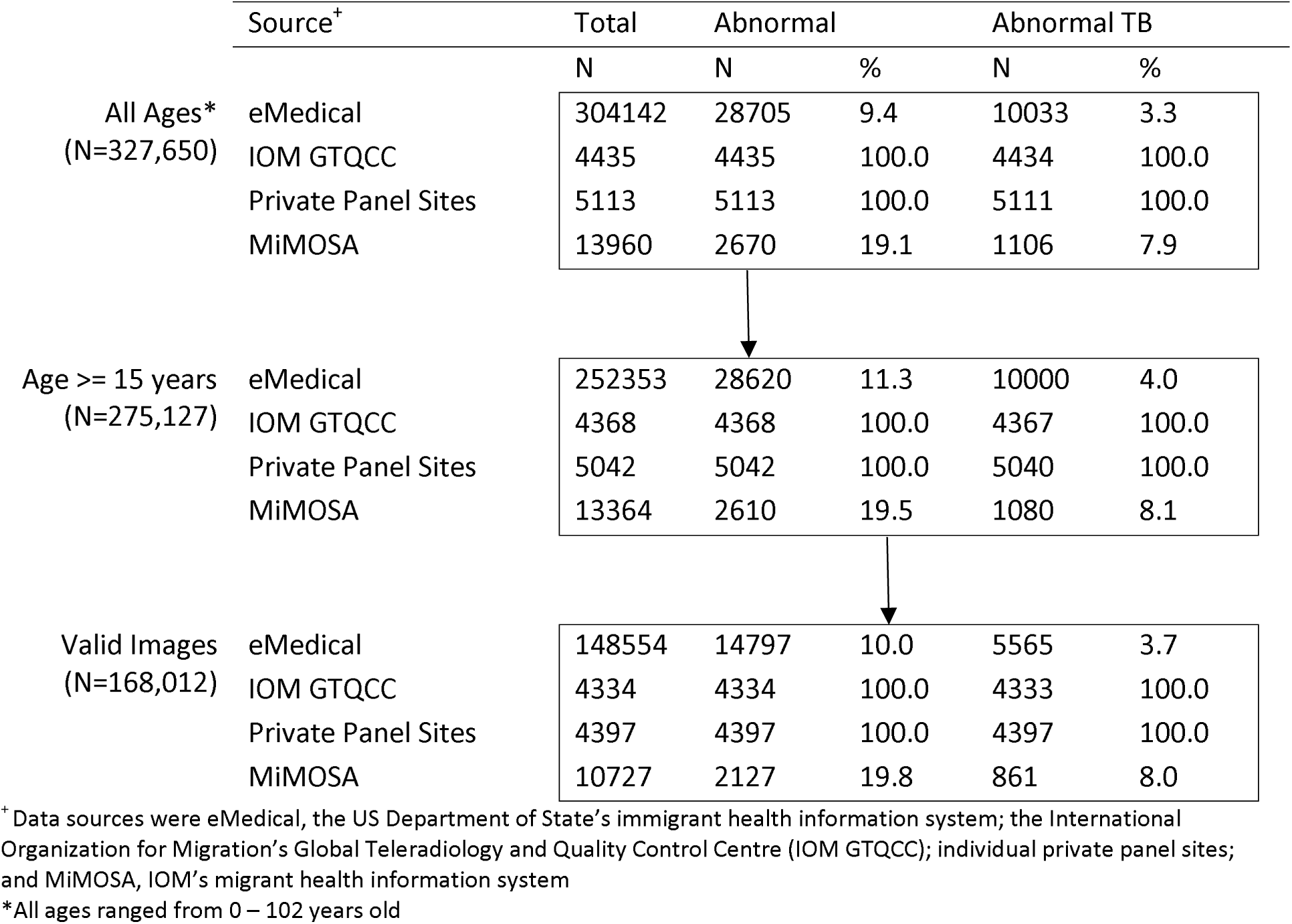
Flow diagram detailing the number of radiographs by data source and abnormality status from before exclusion for age (All Ages), after exclusion for age under 15 years, and after exclusion for whether the DICOM pixel arrays were Python-readable (Valid Images). Radiographs were collected between the years of 2011 and 2021 and constitute a convenience sample of the total applicant population for that time period.

### Radiologist annotations

Chest radiograph abnormalities for the immigration exam fall into one of two groups: those suggestive of tuberculosis and those not. Abnormalities suggestive of tuberculosis include: infiltrates or consolidations; reticular markings suggestive of fibrosis; cavitary lesions; nodules or mass with poorly defined margins; pleural effusions; hilar or mediastinal adenopathy; miliary findings; discrete linear opacities; discrete nodule(s) without calcification; volume loss or retraction; and irregular thick pleural reaction. Abnormalities not suggestive of tuberculosis include cardiac, musculoskeletal, or other abnormalities; smooth pleural thickening; diaphragmatic tenting; calcified pulmonary nodules; and calcified lymph nodes.

Most abnormal images in our internal validation and test sets were suggestive of tuberculosis. Although we did benchmark our generic model against two open datasets with a wider range of abnormalities (described below), we focused primarily on the tuberculosis classification task for our analysis. Importantly, however, because only a small number of the abnormal images (1,551) were from applicants with active tuberculosis at the time of screening—the vast majority were from applicants who had previously been screened, diagnosed with tuberculosis, and received treatment, or whose tuberculosis sputum testing results were negative—we chose not to benchmark our models against a microbiological or bacteriologic reference standard, focusing instead on a purely radiological reference standard.

### External test sets

To supplement our internal testing data, we benchmarked our binary models 1 and 2 on four external datasets: ChestX-ray8^33^; the Montgomery County, USA (MCU) and the Shenzhen, China (SHN) tuberculosis datasets^34^; and VinDr-CXR.^35^ VinDr-CXR was the largest with 2,971 images in total, 161 of which were suggestive of tuberculosis; the others ranged in size from 138 images (MCU) to 810 images (ChestX-ray8). For all datasets, we use the testing splits specified in their original publications, and the original labels, with the exception of ChestX-ray8, for which we use the refined test labels provided by Google^23,27^.

MCU, SHN, and VindDr-CXR had labels indicating the suggested presence of tuberculosis (reference standards varied by dataset and included radiographic, clinical, and laboratory evidence of disease), but only ChestX-ray8 and VinDr-CXR also had labels indicating a variety of other abnormalities. Because of this imperfect overlap between our classification tasks and the labels in the datasets, VinDr-CXR is the only dataset on which we test both binary models (1 and 2); for the other three, we test only Model 1 (ChestX-ray8) or Model 2 (MCU and SHN).

### Dataset splitting

For our internal data, we began with 168,012 images in total, which we then randomly split into training (152,012; 15% abnormal), validation (8,000; 50% abnormal), and testing (8,000; 50% abnormal) sets, following a sample size calculation we used to determine the number of images we would need to achieve a 5% margin of error in estimating sensitivity (technical details on the procedure are provided in the Supplemental Methods). Training images were single-read images randomly drawn from all sites. Testing and validation images for Task 2 had either been double-read as part of the IOM Teleradiology QA/QC program or single-read at a handful of panel sites in areas with high TB burden.

For ChestX-ray8, we reserved an additional 8,000 images from the original training data to serve as validation data for Task 1 (in our internal validation dataset, abnormalities not suggestive of tuberculosis were underrepresented, as the abnormal images were almost always abnormal and suggestive of tuberculosis).

### Operating point selection

When validation data was available, we used it to select two operating points for thresholding the models’ predictions on the corresponding test sets: one that maximized Youden’s J index (all tasks), and one that minimized the relative error in predicted prevalence (Tasks 2 and 3 only). We named these two operating points the “J” and “count” operating points, respectively. Because the proportion of abnormal images in our internal test set was different than the corresponding proportion in the training set, the latter being generally representative of the screening program’s data distribution over a multiyear period of time, the count-based operating points were selected using a reweighting scheme that minimized error in predicting the proportion from the training set using the model’s performance characteristics (sensitivity and specificity) on the validation set; this procedure is described in full in the Supplemental Methods. Finally, when validation data was not available, as was the case for all external datasets except for ChestX-ray8, we selected a single operating point that maximized Youden’s J index on the test sets. We provide all operating points in Supplemental Table 2.

### Image preprocessing

After discarding DICOM files with corrupt pixel arrays, we extracted the pixel arrays and saved them as 1024×1024-pixel PNG files. We then used optical character recognition (OCR) software to identify images with evidence of burned-in patient metadata and removed them from the dataset. We describe both of these procedures more fully in the Supplemental Methods.

### Model architecture and training procedures

To improve the model’s ability to generalize to unseen data, we used a custom image augmentation layer as the input layer, randomly perturbing brightness, contrast, saturation, and other characteristics to the radiographs during training; value ranges for these perturbations were taken from Majkowska et al. 2020 and remained fixed during training.^27^ For the feature extractor, we used EfficientNetV2M^36^, which was pretrained on ImageNet.^37^ The final layers in our model were a dropout layer (probability = 0.5, held fixed) and then a dense layer with a sigmoid activation and binary cross-entropy loss.

We trained all models in minibatches of 12 images (4 per GPU) with the Adam^38^ optimizer and a fixed learning rate of 1e-4. For all tasks, we allowed training to continue until AUC began to decrease on the validation data at which point we saved the model weights and proceeded to testing.

### Statistical inference

We calculated common classification performance metrics for all models and test sets, including AUC, sensitivity, specificity, and F1. For tuberculosis-specific datasets, we also calculated specificity at 90% sensitivity and sensitivity at 70% specificity, in line with the WHO’s TPP for tuberculosis triage tests for use in community settings. For the HaMLET test set, we calculated the model’s relative error in predicting prevalence (i.e., the true number of abnormal-TB images), mirroring our primary operational use-case for the model as a tool for internal QC activities.

For all metrics, we calculated bias-corrected and accelerated (BCA) bootstrap confidence intervals^39,39^ down-sampling abnormal images in the bootstrap replicates so the percentage of abnormal images in each was equal to the percentage in the training data (target percentage for each task provided in Table 1; see the Supplemental Methods for more details). We did not adjust the intervals for multiplicity.

**Table 1.** Distributions of age and sex for the applicants in our training, validation, and testing datasets, along with the geographic distribution of their corresponding health screening exam sites.

|  |  | All |  | Training |  | Validation |  | Testing |  |
| --- | --- | --- | --- | --- | --- | --- | --- | --- | --- |
|  |  | N | % | N | % | N | % | N | % |
|  | Total | 168,012 | -- | 152,012 | 90 | 8000 | 5 | 8000 | 5 |
| Age Group | 15-24 | 37837 | 23 | 34797 | 23 | 1541 | 19 | 1499 | 19 |
|  | 25-34 | 40715 | 24 | 37549 | 25 | 1567 | 20 | 1599 | 20 |
|  | 35-44 | 28614 | 17 | 26035 | 17 | 1268 | 16 | 1311 | 16 |
|  | 45-54 | 25581 | 15 | 23032 | 15 | 1302 | 16 | 1247 | 16 |
|  | 55-64 | 19949 | 12 | 17581 | 12 | 1159 | 14 | 1209 | 15 |
|  | >=65 | 12029 | 7 | 9973 | 7 | 1050 | 13 | 1006 | 13 |
|  | N/A | 3286 | 2 | 3044 | 2 | 113 | 1 | 129 | 2 |
| Sex | F | 92851 | 55 | 86364 | 57 | 3261 | 41 | 3226 | 40 |
|  | M | 70826 | 42 | 65301 | 43 | 2742 | 34 | 2783 | 35 |
|  | N/A | 4334 | 3 | 346 | 0 | 1997 | 25 | 1991 | 25 |
| Data Source<sup>+</sup> | IOM GTQCC | 4334 | 3 | 346 | 0 | 1997 | 25 | 1991 | 25 |
|  | MiMOSA | 10727 | 6 | 10727 | 7 | 0 | 0 | 0 | 0 |
|  | Private Panel Sites | 4397 | 3 | 385 | 0 | 2003 | 25 | 2009 | 25 |
|  | eMedical | 148553 | 88 | 140553 | 92 | 4000 | 50 | 4000 | 50 |
| Exam Region* | Africa | 29753 | 18 | 28854 | 19 | 430 | 5 | 469 | 6 |
|  | Americas | 43296 | 26 | 33707 | 22 | 4803 | 60 | 4786 | 60 |
|  | Asia | 77755 | 46 | 72722 | 48 | 2538 | 32 | 2495 | 31 |
|  | Europe | 15369 | 9 | 14895 | 10 | 228 | 3 | 246 | 3 |
|  | Oceania | 1453 | 1 | 1453 | 1 | 0 | 0 | 0 | 0 |
|  | N/A | 385 | 0 | 380 | 0 | 1 | 0 | 4 | 0 |
| Exam Sub-Region | Australia and New Zealand | 1276 | 1 | 1276 | 1 | 0 | 0 | 0 | 0 |
|  | Central Asia | 996 | 1 | 996 | 1 | 0 | 0 | 0 | 0 |
|  | Eastern Asia | 11634 | 7 | 11634 | 8 | 0 | 0 | 0 | 0 |
|  | Eastern Europe | 7884 | 5 | 7410 | 5 | 228 | 3 | 246 | 3 |
|  | Latin America and the Caribbean | 38336 | 23 | 28747 | 19 | 4803 | 60 | 4786 | 60 |
|  | Melanesia | 172 | 0 | 172 | 0 | 0 | 0 | 0 | 0 |
|  | Northern Africa | 4883 | 3 | 4882 | 3 | 0 | 0 | 1 | 0 |
|  | Northern America | 4960 | 3 | 4960 | 3 | 0 | 0 | 0 | 0 |
|  | Northern Europe | 1204 | 1 | 1204 | 1 | 0 | 0 | 0 | 0 |
|  | Polynesia | 5 | 0 | 5 | 0 | 0 | 0 | 0 | 0 |
|  | South-eastern Asia | 34368 | 20 | 30552 | 20 | 1912 | 24 | 1904 | 24 |
|  | Southern Asia | 21561 | 13 | 20452 | 13 | 564 | 7 | 545 | 7 |
|  | Southern Europe | 5020 | 3 | 5020 | 3 | 0 | 0 | 0 | 0 |
|  | Sub-Saharan Africa | 24870 | 15 | 23972 | 16 | 430 | 5 | 468 | 6 |
|  | Western Asia | 9196 | 5 | 9088 | 6 | 62 | 1 | 46 | 1 |
|  | Western Europe | 1261 | 1 | 1261 | 1 | 0 | 0 | 0 | 0 |
|  | N/A | 385 | 0 | 380 | 0 | 1 | 0 | 4 | 0 |

+IOM GlobalGlobal Teleradiology and Quality Control Center (GTQCC) and the private panel sites all have their own internal quality control processes.
\*Exam Region:
**Africa:** Algeria, Angola, Benin, Burkina Faso, Burundi, Cameroon, Cape Verde, Chad, Democratic Republic of the Congo, Republic of the Congo, Djibouti, Egypt, Gambia, Ghana, Guinea, Ivory Coast, Kenya, Liberia, Malawi, Morocco, Mozambique, Namibia, Niger, Nigeria, Rwanda, Senegal, South Africa, Sudan, Tanzania, Togo, Tunisia, Uganda, Zambia, and Zimbabwe
**Americas:** Argentina, Barbados, Belize, Bolivia, Brazil, Canada, Chile, Colombia, Costa Rica, Dominican Republic, Ecuador, Guatemala, Guyana, Haiti, Honduras, Jamaica, Mexico, Nicaragua, Panama, Paraguay, Peru, Trinidad and Tobago, and Uruguay
**Asia:** Afghanistan, Azerbaijan, Bahrain, Bangladesh, Cambodia, China, Hong Kong, India, Indonesia, Iraq, Israel, Japan, Jordan, Kazakhstan, Republic of Korea, Kuwait, Kyrgyzstan, Laos, Lebanon, Malaysia, Mongolia, Myanmar, Nepal, Oman, Pakistan, Philippines, Saudi Arabia, Singapore, Sri Lanka, Taiwan, Tajikistan, Thailand, Turkey, Turkmenistan, United Arab Emirates, Uzbekistan, Vietnam, and West Bank
**Europe:** Albania, Austria, Belarus, Belgium, Bosnia and Herzegovina, Bulgaria, Croatia, Cyprus, Denmark, Estonia, Finland, France, Georgia, Germany, Greece, Hungary, Italy, Kosovo, Latvia, Lithuania, Malta, Moldova, Netherlands, North Macedonia, Norway, Poland, Portugal, Romania, Russia, Serbia, Slovak Republic, Slovenia, Spain, Sweden, Switzerland, Ukraine, and United Kingdom
**Oceania:** Australia, Fiji, New Zealand, Papua New Guinea, and Tonga

### Abnormality localization

We used two saliency methods, Grad-CAM^40^ and XRAI^41^, to generate abnormality heatmaps for the images. We examined a small selection of the heatmaps for true-positive and false-positive images (abnormal and normal images, respectively, with high model-based probabilities of abnormality) to explore their use as approximate abnormality localization methods. Because we did not have ground-truth bounding box annotation for the images, this step was primarily exploratory.

### Software and Hardware

Our code is publicly available at https://github.com/cdcai/hamlet.git. Complete information on the software and hardware used is available in the Supplemental Methods.

### Ethical considerations

This project was proposed, reviewed, and approved in accordance with CDC institutional review policies and procedures. Because it received a non-research determination, review by an institutional review board was not required. Neither trained model weights nor raw images will be made publicly available to protect applicant privacy.

## Results

### Demographic characteristics of our study sample

Table 1 shows the demographic characteristics of the applicants in our study sample, and where their screening exams were conducted. Overall, 55% of the applicants were women, and most (64%) were between the ages of 15 and 44. Applicants 65 years and older were the rarest, constituting 7% of both the overall and training data, followed by applicants between 55 and 64 (12%) and those between 45 and 54 (15%). In our validation and test sets, applicants in the youngest age group (15 to 24) were underrepresented (19% in each), while those in the oldest age group (65 and over) were overrepresented (13% in each), relative to the age distribution in both the overall sample and the training data.

Geographically, most of the exams were conducted in Asia (46%), with Southeastern Asia (20%), Southern Asia (13%), and Eastern Asia (7%) being the three primary contributors to the region. By sub-region, most exams were conducted in Latin America and the Caribbean (23%), which was the main contributor to exam volume in the Americas region (26% of exams overall). By contrast, Oceania (1%) and Europe (9%) had the smallest representation by volume. In our validation and test datasets, these percentages changed substantially, with Latin America and the Caribbean contributing 60% of the images to both, and Southeastern and Southern Asia contributing 31%, with the remaining 9% comprising images from Africa (6%) and Europe (3%).

By data source, the majority of our images (88%) came from eMedical, the US Department of State’s immigrant health data system. Of the remaining images, 6% came from MiMOSA, IOM’s health data system, 3% from the IOM Teleradiology QC program, and 3% from our partner panel sites. Although the training dataset skewed heavily toward images from eMedical (92%), the validation and test datasets were evenly split between eMedical (50%) and the IOM Teleradiology QC program (25%) and our partner panels (25%). As mentioned above, the latter two primarily contributed images that were abnormal and suggestive of TB, and so we used eMedical as the source for normal images, noting that these latter images were also drawn from screenings performed by our partner panel sites and not from the system at random.

### Distribution of abnormalities

Table 2 shows the distribution of general and specific findings across our internal training, validation, and test sets. In the training data, 12% of the images were abnormal and 5% were abnormal and suggestive of tuberculosis with 0.1% of images in the latter category from applicants who were either smear-or culture-positive for tuberculosis disease at the time of screening. In the validation and test sets, these percentages changed to 50%, 50%, and 9% respectively, both because we up-sampled abnormal images to increase precision in estimating sensitivity, and because we had requested additional images from smear-or culture-positive applicants from the IOM Teleradiology QC program and our partner panel sites to use for testing.

**Table 2.**
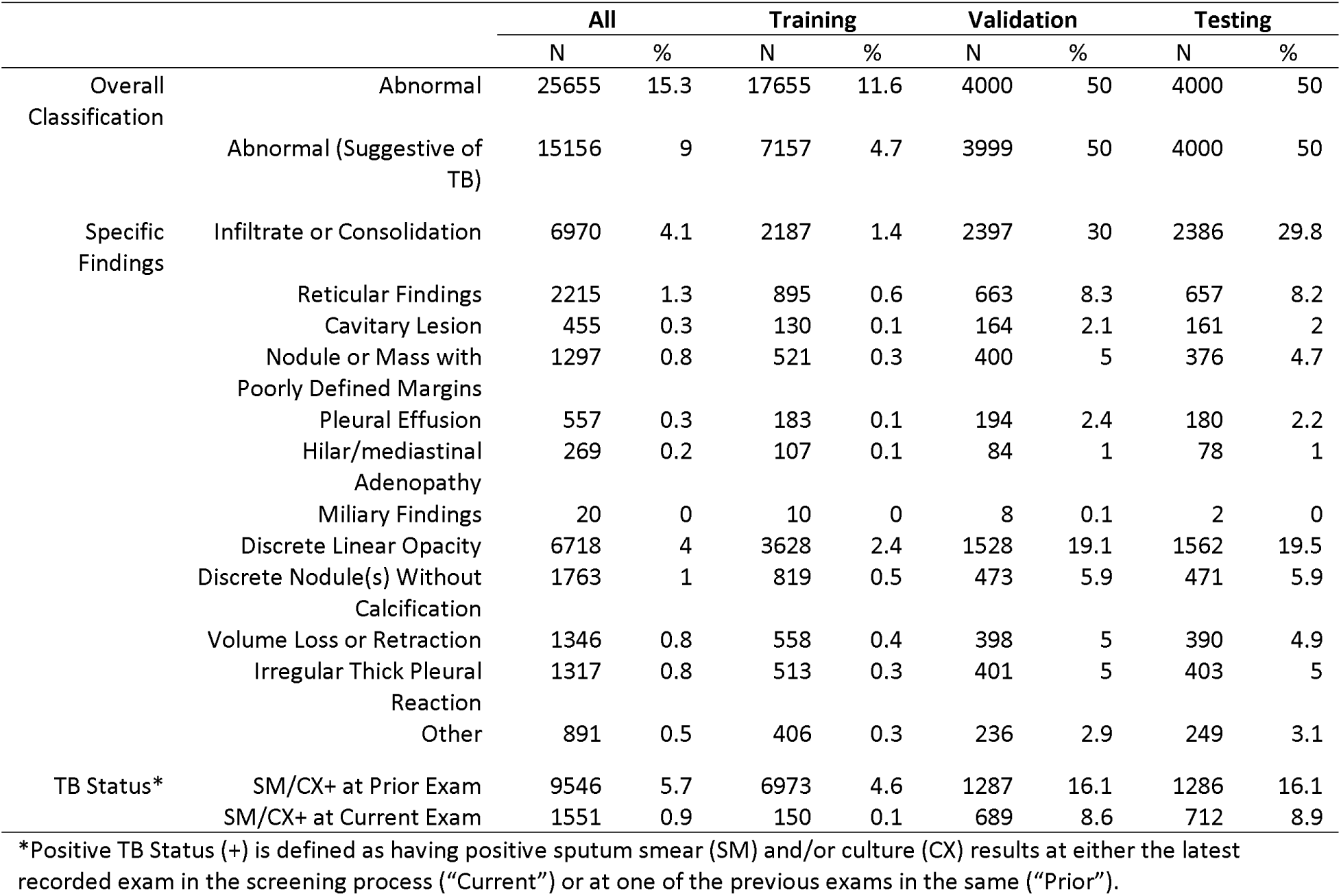
Counts and proportions of radiographic findings suggestive of tuberculosis (TB) in our internal training, validation, and testing datasets.

By specific finding, the most common abnormalities were the discrete linear opacity (2.4% in training; 20% in validation; 20% in testing) and the infiltrate or consolidation (1.4%; 30%; 30%). In the training data, the rarest abnormalities were miliary findings (<1%), cavitary lesions (0.1%), pleural effusions (0.1%), and hilar/mediastinal adenopathy (0.1%), all of which remained rare in the validation and testing data, despite the up-sampling of abnormal images.

### Binary classification performance

Table 3 shows the performance metrics for our models on the two binary classification tasks. For Task 2, AUCs were consistently high, ranging from 0.99 (95% CI 0.97, 1.0) on MCU to 0.94 (0.93, 0.95) on VinDr-CXR. Specificity at a sensitivity of 0.90 was similarly high, ranging from 0.83 (0.78, 0.87) on VinDr-CXR to 0.98 (0.89, 1.0) on MCU, although for reasons we provide in the discussion, we do not suggest whether any of these would meet the WHO’s TPP for tuberculosis triage tests. With the count-based operating point, the model also fared well in predicting the number of abnormal images suggestive of tuberculosis in our internal test set, achieving a relative error of only −2% (−8%, 6%).

**Table 3.**
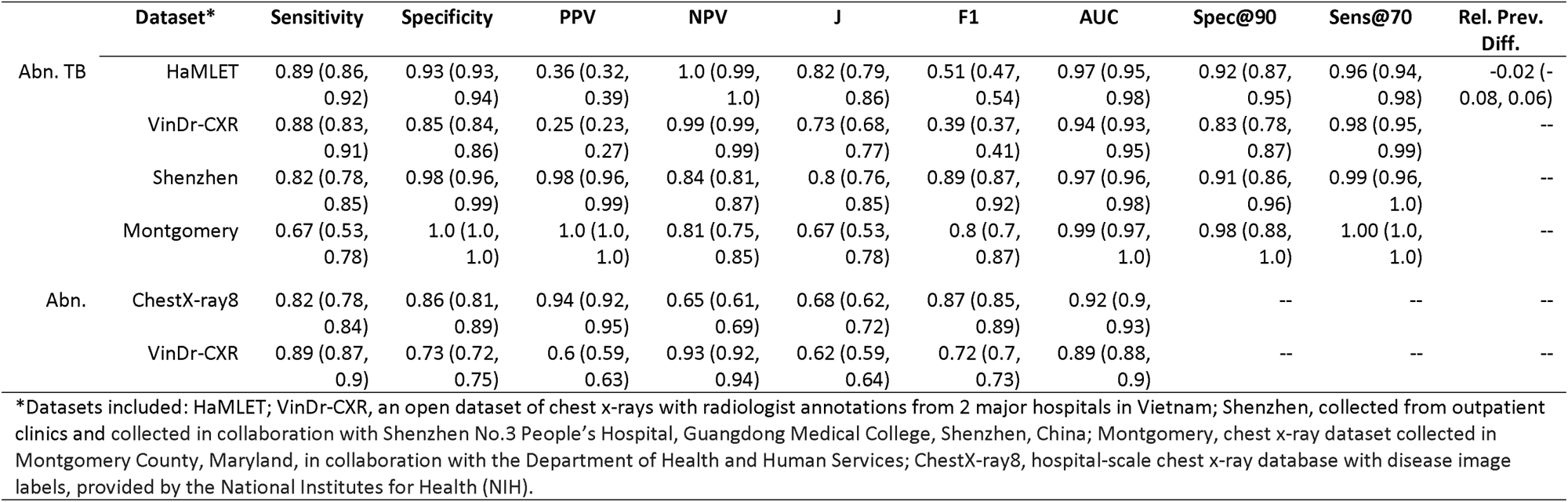
Classification results for our abnormal-TB and abnormal models on our internal dataset and on four external datasets.

Performance was similar, although slightly lower, on Task 1. On ChestX-ray8, the model achieved an AUC of 0.92 (0.90, 0.93) and an optimal sensitivity and specificity of 0.82 (0.78, 0.84) and 0.86 (0.81, 0.89); on VinDr-CXR, these numbers were 0.89 (0.88, 0.90), 0.89, (0.87, 0.90), and 0.73 (0.72, 75), respectively.

Because we did not have internal testing data for this task, we did not test this model with the count-based operating point.

### Multilabel classification performance

Table 4 shows the performance metrics for our models on Task 3. AUCs ranged from 0.78 (95% CI 0.73, 0.8) in predicting discrete linear opacities to 0.96 (0.65, 0.99) in predicting pleural effusions. With the J operating point, sensitivity and specificity jointly reached 0.8 for four of the abnormalities: infiltrates, cavities, pleural effusions, and volume loss or retraction. For the other abnormalities, sensitivity at that operating point was higher than specificity, sometimes by a large margin (e.g., 0.91 sensitivity and 0.53 specificity for discrete nodules without classification). Because most abnormalities were rare in our data, metrics that depend on prevalence, like PPV and F1, were consistently low, with F1 reaching a maximum of 0.13 (0.10, 0.15) for infiltrates and dipping as low as 0.00 (0.00, 0.01) for hilar adenopathies.

**Table 4.**
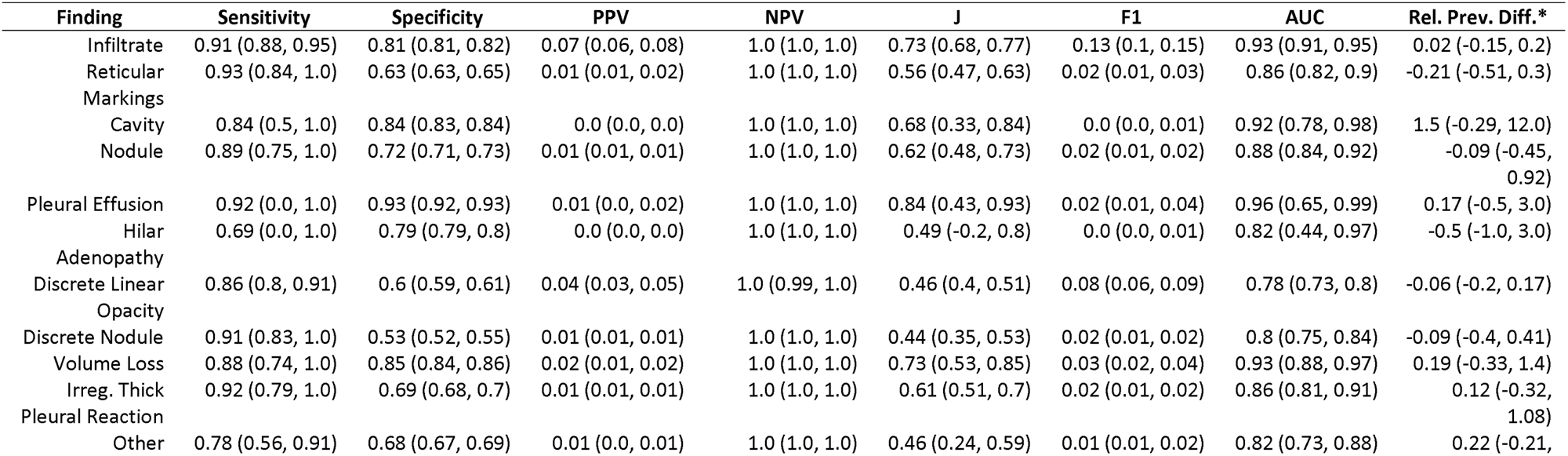
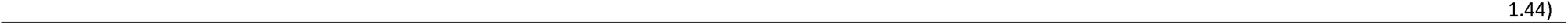
Classification metrics for our findings-specific model on our internal dataset.

With the count operating point, the model produced fairly accurate prevalence estimates for a number of the abnormalities, with three absolute relative errors under 10% (2% for infiltrates, −6% for discrete linear opacities, and −9% for both kinds of nodules), and three under 20% (12% for irregular thick pleural reactions, 17% for pleural effusions, and 19% for volume loss or retraction). We note here that operating point selection is crucial for accuracy on this task with relative errors rising by several orders of magnitude when the J operating point was used instead of the count operating point (Supplemental Table 2).

### Approximate abnormality localization

Figure 2 shows Grad-CAM (second and third columns) and XRAI (fourth and fifth columns) for five radiographs correctly identified as abnormal by Model 1; the radiographs were drawn from the abnormal examples in ChestX-ray8, SHN, and VinDR-CXR. Figure 3 shows the same panel of heatmaps, but for five images incorrectly identified as abnormal, also by Model 1 and drawn from the same datasets. In general, the two methods identify similar regions of the radiographs as being abnormal, although they do occasionally diverge in both the extent and the severity of the highlighted regions.

**Figure 2.**
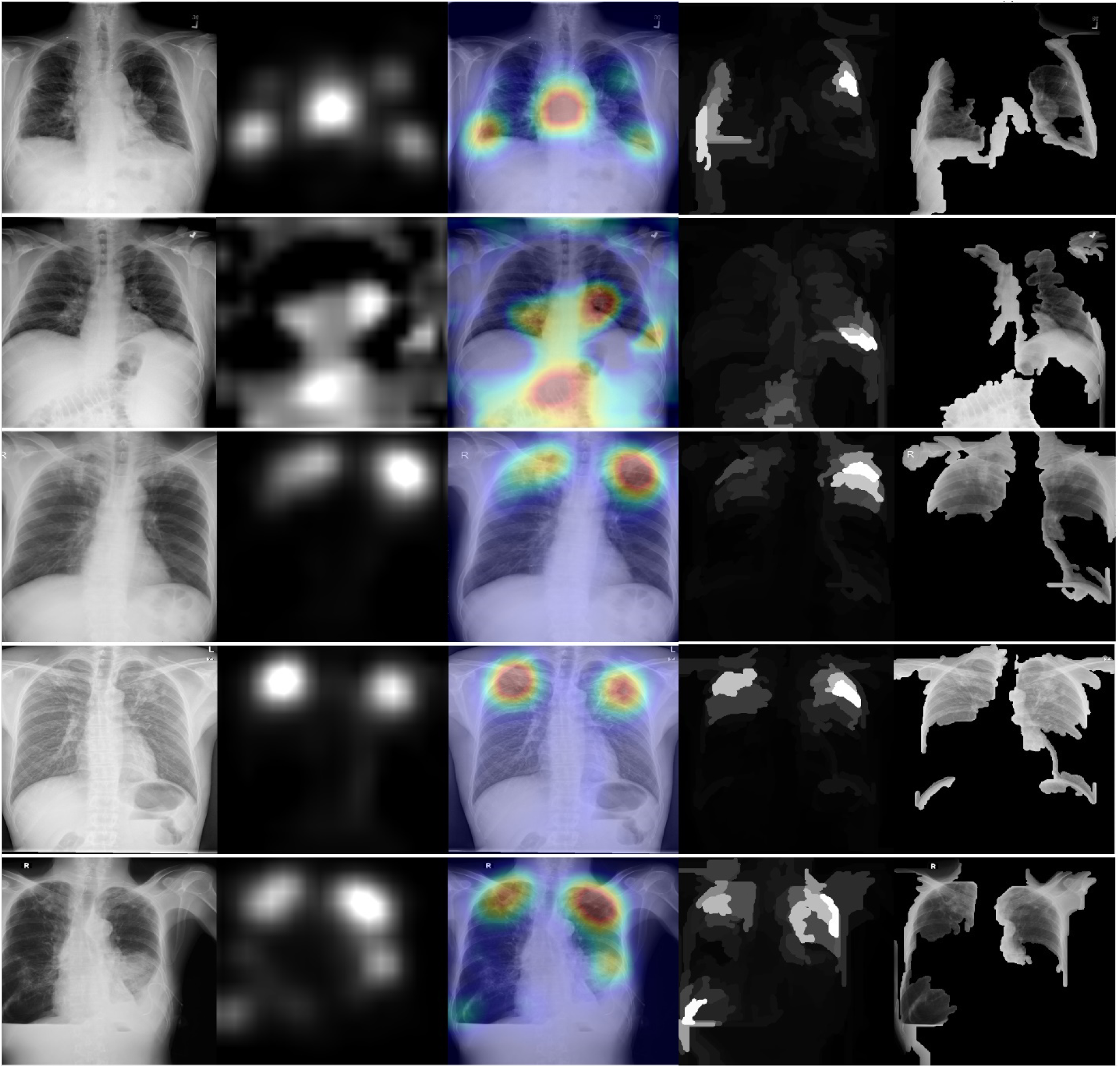
XRAI (left) and GradCAM (right) heatmaps for true positive images. The original radiographs are on the left, the Grad-CAM activations and heatmaps are in the middle, and the XRAI activations and overlays are on the right. For the XRAI overlays, only regions reaching the 70^th^ percentile of activation strength are shown.

**Figure 3.**
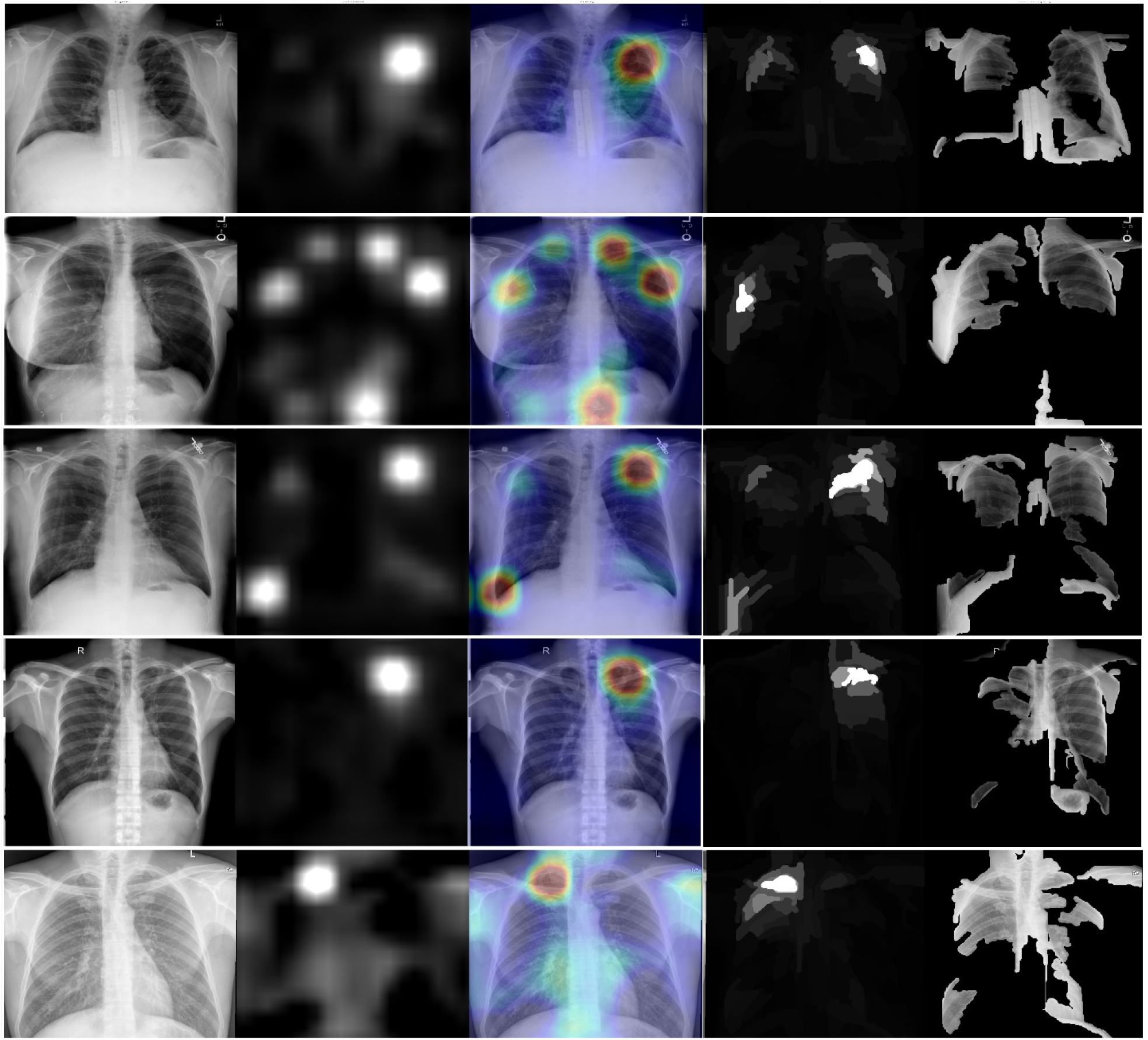
XRAI (left) and GradCAM (right) heatmaps for false positive images. The original radiographs are on the left, the Grad-CAM activations and heatmaps are in the middle, and the XRAI activations and overlays are on the right. For the XRAI overlays, only regions reaching the 70^th^ percentile of activation strength are shown.

## Discussion

### Applications of current models

Our binary models showed strong results on our internal datasets at both of our chosen operating points. Although we do not currently intend for either model to be used for individual-level classification tasks, e.g., as part of the overseas immigration exam clinical workflow, their performance is on-par with top-performing models published in the literature, including several commercial products designed largely to detect radiographic signs of tuberculosis (see Codlin 2021 and Kik 2022).^42,43^ Performance on the external datasets also suggests that the models may generalize well to unseen data, even on tasks for which we did not have clean validation or testing data, like Task 1, on which our models come within two percentage points of what we believe to be the current state-of-the-art on AUC (0.92 from Model 1 vs. 0.94 from Nabulsi et al. 2021).^23^ The main exception to this trend is Model 3, which despite achieving good AUCs in identifying many of the specific findings, is not likely to be clinically useful, at least in the population represented by our internal data, owing to the rarity of most of the findings and the model’s resulting poor PPV.

A relatively clear use-case for our tuberculosis-specific models (2 and 3), however, is in estimating sample-level counts of abnormal images and, depending on the abnormality in question, specific findings. As a tool for conducting internal QC on radiographs read during the overseas immigration exam, we can imagine running the models on batches of incoming images and comparing their number of positive calls to the number of reported abnormal radiographs, raising an alert when the difference in counts exceeds a predefined threshold and triggering a model-guided manual review for further investigation. In the case of Model 2, for example, which underestimated the number of abnormal images suggestive of tuberculosis in our internal test set by only 2%, a reasonable threshold might be +/− 10%, just beyond the bounds of the 95% CI (−8% to 6%), and the corresponding manual review might begin with the images with the highest model-based probability of abnormality among those initially reported normal. This kind of process may be stratified by key operational variables, like exam site or country, and may also be informed by existing epidemiologic information, like the expected background rates of tuberculosis disease in the screening areas or site-specific historical rates of abnormal radiographs confirmed by earlier QC efforts.

### Directions for future research

In our case, a natural first step for future research would be a follow-up validation study with manual review to explore our models’ utility tools for supporting internal QC efforts. A number of studies have examined the performance of already-trained models, mostly in the form of commercial software, in detecting abnormalities^43^ and, in certain cases, tuberculosis disease (Khan 2020; Qin 2021).^26,30^ To our knowledge, ours is the first study to propose evaluating models’ ability to estimate sample-level counts, and more evaluation would be needed before integrating them with existing QC workflows. A designed QC study would also allow for the evaluation of attribution methods, like the saliency heatmaps we produced for Figures 2 and 3, as tools for abnormality localization to assist with manual review, which only a small number of prior studies have rigorously addressed.^44,45^

Similarly, an operational analysis to decide when, where, and how to use the models to improve screening programs would fill a gap in the literature, which to date has focused primarily on examining model performance in clinical contexts rather than the downstream effects of incorporating them into larger workflows. Remaining problems include establishing best practices for selecting operating points by country or facility to optimize detection given local constraints on resources and background rates of tuberculosis; estimating minimum diagnostic performance needed to achieve cost-effectiveness and programmatic efficiency under different operating scenarios; and evaluating the epidemiologic and economic impact of allowing radiologists to use the models for decision support during the overseas screening exams. There is evidence that similar models can improve turnaround time^23^ or lower costs^29^ associated with diagnostic workflows, and because of the scope of the CDC’s overseas screening program, these seem like potentially fruitful avenues of investigation.

A final direction for future research is in developing and evaluating models for predicting active tuberculosis disease from chest radiographs in combination with relevant clinical, demographic, and immunologic information. To our knowledge, no model in the literature has been trained from the start to predict tuberculosis disease from the radiograph directly, although training datasets may use a mix of radiographic, clinical, and microbiological results as reference standards; and no model has been developed that accepts multimodal (i.e., non-radiographic) inputs. Given these limitations, performance in predicting tuberculosis disease is determined primarily by two pieces of information: a model’s performance in identifying images with abnormalities, and the correlation between the presence of those abnormalities and tuberculosis disease in the target population. Occasionally the correlation is high enough for models to meet the TPP, but often it is not, with observed specificities at 90% sensitivity ranging from well over 70%^26^ to 60% and below^25^ in several well-known commercial algorithms. Using other available information for prediction, whether by stacking an additional model on top of the image-processing module or by altering the module itself to accept multimodal inputs, may yield diagnostic gains, not only because the approach mirrors the process by which clinical diagnoses are made and thus seems *a priori* sensible, but also because patient characteristics like HIV status and history of tuberculosis are known to affect model performance based on the radiographs alone.^26,46^ Although some technical innovation may be required to make this approach feasible, it may well improve our ability to predict tuberculosis disease, especially in low-resourced settings where access to trained radiologists is limited, and thus seems well worth pursuing.

## Conclusion

Using data collected from immigrants and refugees during overseas immigration exams prior to entry into the US, we trained and tested three deep learning models for identifying abnormalities on chest radiographs. The models performed well, achieving high scores on our internal test dataset, and nearing state-of-the-art on several external test datasets.

## Data Availability

Due to the sensitive nature of the populations under study, and to ensure applicant privacy, neither trained model weights nor raw images will be made publicly available.

https://github.com/cdcai/hamlet

## Abbreviations

AI: artificial intelligence
AP: anterior-posterior
AUC: area under the ROC curve
CDC: Centers for Disease Control and Prevention
M
CNN: convolutional neural network
DGMH: Division of Global Migration Health
HaMLET: Harnessing Machine Learning to Eliminate Tuberculosis
IOM: International Organization for Migration
IRHB: Immigrant and Refugee Health Branch
MiMOSA: Migrant Management Operational System Application
MTB: *Mycobacterium tuberculosis*
NLP: natural language processing
NAAT: nucleic acid amplification testing
OCR: optical character recognition
PII: personally-identifiable information
PPV: positive predictive value
QC: quality control
ROC: receiver operating characteristic
TPP: target product profile
WHO: World Health Organization

## Acknowledgements

The authors thank the following panel sites and organizations for their contribution to this work: Clinica Medica Internacional (Mexico), Servicios Medicos Consulares (Dominican Republic), Servicios Medicos de la Frontera (Mexico), and St. Luke’s Medical Center Extension Clinic (the Philippines), IOM Telerad QC, and the International Panel Physician Association. We also thank colleagues for review and support as well as Mary Naughton, Jessica Webster, Nina Marano,, Sifrash Gelaw, Bhaskar Amatya, Alexandra Ortega, and Zachary White.

## SUPPLEMENT

### Additional Information on the Overseas Screening Procedures

X- ray images collected during the overseas health screenings are interpreted by a radiologist at the panel physician’s office (panel site) and reviewed by the panel physician. The process is to first determine if the chest x-ray image is normal or abnormal, if abnormal then the radiologist will determine if the image suggests that the applicant has tuberculosis (TB). If the image is suggestive of TB, then the applicant needs sputum smears and culture testing done. For the images that suggest the applicant has TB, the radiologist identifies and records the specific radiographic features associated with this determination, and the panel physician then determines each applicant’s TB classification and records that information on the applicant’s US Department of State TB Worksheet (DS-3030 form).

Any applicant diagnosed with pulmonary TB disease must receive a classification of Class A TB and is not cleared for travel until successfully finishing directly observed treatment under the supervision of a panel physician. Those who receive a classification of Class B0 TB (applicants, previously classified as Class A, who successfully completed treatment), Class B1 TB (applicants with signs or symptoms or chest x-ray findings suggestive of TB disease, or known HIV infection, but negative sputum smears and cultures and are not diagnosed with TB disease), Class B2 TB (applicants with a positive IGRA, but otherwise have a negative evaluation for TB; i.e., latent TB infection), or Class B3 TB (applicants who are a recent contact of a known TB disease case, regardless of IGRA or TST results) are cleared and permitted to travel.

### Supplemental Methods

#### Sample size calculation

We used a simulation procedure to determine the number of abnormal images we would need in our validation and test sets to accurately estimate sensitivity, specificity, and other key performance metrics for our models. As a starting point, we assumed the models would achieve a sensitivity and specificity of 80% for the binary classification tasks, which is roughly in line with similar models in the literature (see main manuscript for examples). The estimation procedure itself followed these steps:

1. Choose a sample size *N_s_*. In our simulation, these ranged from 100 to 15,000.
2. Generate a single random variate *k_abn_* from a binomial distribution with *n* = *N*_s_ and *p* = *p*_abn_, where *p*_abn_ is the expected prevalence of abnormal images suggestive of TB (in our case, this was 7%).
3. Generate random variates for true positives (TP), false positives (FP), false negatives (FN), and true negatives (TN) with a single draw from a multinomial distribution, where *n* = *N_s_* and *p_k_* is the probability of success for each category *k* as determined by the prespecified levels *p*_se_ and *p*_sp_ of sensitivity and specificity, respectively. Explicitly, these are:

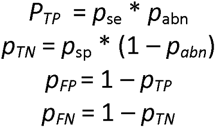
4. Repeat steps 1 and 2 a large number of times (in our case 1,000) and record the full spectrum of performance metrics based on the resulting confusion matrices.
5. Record the 2.5^th^ and 97.5^th^ percentiles for each metric, and take the half-width of the resulting interval as the statistical precision in estimating the metric at the given sample size *N*_s_.
6. Increase sample size by 100 and repeat steps 2 through 6 until the maximum sample size has been exceeded.

We aimed for a minimum sample size that would allow us to calculate sensitivity and specificity to within 5%. Both our validation and test sets exceeded this minimum by a large margin with an approximate precision for each metric of 1.25%.

#### Image preprocessing

We began by excluding from our datasets radiographs with corrupt DICOM files or corrupt pixel arrays. For DICOM files without such corruptions, we then extracted their raw pixel arrays, converted them to grayscale, and exported them to 1,024×1,024-pixel PNG files. Once the image files were saved, we ran a separate script that used optical character recognition (OCR) software (detailed in *Software and hardware* below) to identify radiographs with burned-in annotations and discarded any identified as containing more than three words of text. This exclusion step served mainly to protect the identities of the entrants by removing personally identifiable information (PII) from the images, but it also prevented the text information from affecting the model’s performance. Finally, we discarded images with mean pixel values (out of 255) below 50 or above 215 which were either too dark or too light, respectively, for radiographic features to be visible.

#### Model architecture

To improve the model’s ability to generalize to unseen data, we used a custom image augmentation layer as the input layer with the following transformations and deltas:

Horizontal flips (0.5 probability);
Resizing (either 0.7x, 0.85x, 1.15x, or 1.3x the original dimensions);
Changes in brightness (delta range −0.525 to 0.525);
Changes in contrast (delta range 0.349 to 1.346);
Changes in saturation (delta range 0.382 to 1.403); and
Changes in hue (delta range −0.127 to 0.127).

The delta ranges were taken from Majkowska 2020 and did not vary during training, and the transformations were turned off during validation and testing. The (sometimes perturbed) images were then passed to the EfficientNetV2M feature extractor for processing, after which they passed through a dropout layer (p=0.5) and then a randomly initialized final dense layer with either binary or categorical cross-entropy loss, depending on the task, for final classification.

#### Model training and validation procedures

We trained all models with a minibatch size of 12 (4 per GPU), which allowed us to fine-tune all blocks of the feature extractor at once (in larger minibatches, or on GPUs with lower amounts of memory, the blocks may need to be unfrozen sequentially to prevent out-of-memory errors). We trained each model until its AUC on the validation data began to decrease; in our case, this yielded one epoch of training for the abnormal/normal model (Model 1), two for the abnormal-TB model (Model 2), and two for the multilabel model (Model 3). In all cases, we used the Adam optimizer (Kingma 2014) with a fixed learning rate of 0.001.

#### Operating point selection

When validation data was available, we used it to select two operating points for thresholding a model’s predictions on the test data: one that maximized Youden’s J index and one that minimized the model’s error in predicting the positive samples. We calculated the first directly from the model’s predicted probabilities on the validation data, but we calculated the second by reweighting the positive samples so that test data prevalence equaled the prevalence in the entire dataset before splitting (they were up-weighted in the validation and test sets to improve precision in estimating sensitivity). Our reweighting method was informed by the following observations:

1. Sensitivity (true positive rate, or TPR) and specificity (1 minus the false positive rate, or FPR) are conditional on the true label (e.g., abnormality or the presence of a specific abnormality) and thus independent of prevalence.
2. True prevalence and predicted prevalence are equal when the false positive rate (FPR) and false negative rate (FNR) are equal.
3. When FPR and FNR are equal, the number of counts in the off-diagonal cells of the confusion matrix are equal (this is the mathematical insight behind McNemar’s test for the difference in paired proportions).
4. The off-diagonal cells are equal to the expressions FPR * (1 – *p*) and FNR * *p*, respectively, where *p* is equal to true prevalence.
5. FNR is equal to 1 – sensitivity, and FPR is equal to 1 – specificity.
6. The ROC curve for a given classifier on a test set contains all possible pairs of sensitivity and specificity.
7. The relative difference in prevalence for any given tuple of TPR, FPR, and *p* is given by the expression |[FPR * (1 – *p*)] – [FNR * *p*]| / *p*, where FPR = 1 – specificity and FNR = 1 – sensitivity.

To select the second operating point, then, we simply used the FPR, TPR pairs from a model’s ROC curve on the validation data to calculate the difference between true prevalence and predicted prevalence given a target true prevalence *p_t_*for each possible operating point (using the formula from observation 4 above) and selected the one with the smallest difference.

#### Software and hardware

All models were built with Keras using the TensorFlow 2 backend. DICOM work was done with the pydicom Python package; image preprocessing with the pytesseract, scikit-image, and NumPy packages; data visualization with the seaborn package; and statistical analysis with custom functions for bootstrap resampling. All code is publicly available at https://github.com/cdcai/hamlet.git, and top-level scripts are designed to be rerun on different datasets to aid replication of our results and future research efforts.

Models were trained on a scientific workstation with 32 logical processors, 128GB of RAM, and 3 NVIDIA RTX A6000 GPUs.

#### Bootstrap resampling procedures

To create confidence intervals for the performance metrics, we used the nonparametric bootstrap with bias correction and acceleration (BCA), following Efron 1987. Because the test set contained an even number (4,000) of normal and abnormal images each (we up-sampled abnormal images to increase precision in estimating AUROC and sensitivity), naively constructing the intervals would yield inflated estimates for metrics like positive predictive value (PPV), F1-score, and the relative difference in prevalence that are affected by the underlying prevalence of the abnormal images. To account for this inflation, we constructed each bootstrap replicate by sampling n_ab_ abnormal images from the test set, where n_ab_ is a random draw from a binomial distribution B(n, p), n is the total number of images in the test set (8,000), and p is the proportion of abnormal images in the training data. Each replicate was then filled out with 1 – n_ab_ normal images, also sampled from the test data with replacement, and then the replicate was used to calculate the full range of performance metrics, including sensitivity and specificity (although these are not affected by prevalence). We used this sampling distribution to generate the bias correction parameters z0 and acceleration parameters for the metrics and construct the resulting 95% confidence intervals. As noted in the main manuscript, we did not adjust the intervals for multiplicity.

### Supplemental Results

**Supplemental Table 1.** Operating points for our internal validation set that optimized three criteria: Youden’s J index (j); the absolute relative error in predicted counts (count); and the same relative but reweighted to take the difference in prevalence for each finding between the validation set and the total available data into account (count_adj). We used only the first and third operating points in our analysis.

| Finding | j | count | count_adj |
| --- | --- | --- | --- |
| abnormal | 0.15459 | 0.147612 | 0.448105 |
| abnormal_tb | 0.056503 | 0.037703 | 0.355039 |
| infiltrate | 0.042922 | 0.317821 | 0.860177 |
| reticular | 0.020081 | 0.381927 | 0.530253 |
| cavity | 0.098012 | 0.358573 | 0.773157 |
| nodule | 0.063748 | 0.388444 | 0.890132 |
| pleural_effusion | 0.056165 | 0.736675 | 0.998791 |
| hilar_adenopathy | 0.024143 | 0.328604 | 0.922602 |
| linear_opacity | 0.036351 | 0.371436 | 0.612073 |
| discrete_nodule | 0.008291 | 0.366458 | 0.754047 |
| volume_loss | 0.052816 | 0.305774 | 0.853881 |
| pleural_reaction | 0.028239 | 0.298953 | 0.809463 |
| other | 0.010258 | 0.203865 | 0.672526 |

**Supplemental Table 2.** Distribution of health screenings by year and region. Row and column totals are on the margins.

|  |  | Exam Region |  |  |  |  |  |  |
| --- | --- | --- | --- | --- | --- | --- | --- | --- |
|  |  | Africa | Americas | Asia | Europe | Oceania | N/A |  |
| Exam Year | 2011 | 0 | 3 | 0 | 0 | 0 | 0 | 3 |
|  | 2012 | 0 | 26 | 0 | 0 | 0 | 0 | 26 |
|  | 2013 | 0 | 75 | 170 | 0 | 0 | 0 | 245 |
|  | 2014 | 0 | 27 | 275 | 0 | 0 | 0 | 302 |
|  | 2015 | 66 | 54 | 774 | 79 | 0 | 0 | 973 |
|  | 2016 | 217 | 91 | 846 | 88 | 0 | 2 | 1244 |
|  | 2017 | 173 | 127 | 928 | 83 | 0 | 3 | 1314 |
|  | 2018 | 709 | 694 | 1388 | 196 | 58 | 0 | 3045 |
|  | 2019 | 8826 | 8794 | 18298 | 4038 | 349 | 78 | 40383 |
|  | 2020 | 10010 | 15246 | 31419 | 6006 | 613 | 186 | 63480 |
|  | 2021 | 9752 | 18159 | 23657 | 4879 | 433 | 116 | 56996 |
|  |  | 29753 | 43296 | 77755 | 15369 | 1453 | 385 |  |

## References

1. World Health Organization. Tuberculosis. https://www.who.int/news-room/fact-sheets/detail/tuberculosis Date: 2023 Date accessed: April 21, 2023

2. Centers for Disease Control and Prevention. Latent TB Infection and TB Disease. https://www.cdc.gov/tb/topic/basics/tbinfectiondisease.htm Date: 2020 Date accessed: April 21, 2023

3. Centers for Disease Control and Prevention. Reported Tuberculosis in the United States, 2022. https://www.cdc.gov/tb/statistics/reports/2022/national_data.htm Date: 2021 Date accessed: April 21,2023

4. Centers for Disease Control and Prevention. Technical Instructions for Panel Physicians. https://www.cdc.gov/immigrantrefugeehealth/panel-physicians.html Date: 2021 Date accessed: December 19, 2022

5. Centers for Disease Control and Prevention. Immigrant and Refugee Health. https://www.cdc.gov/immigrantrefugeehealth/index.html Date: 2022 Date accessed: December 19, 2022

6. Liu Y, Posey DL, Cetron MS, Painter JA. Effect of a culture-based screening algorithm on tuberculosis incidence in immigrants and refugees bound for the United States: a population-based cross-sectional study. Ann Intern Med. 2015;162(6):420–428. doi:10.7326/M14-2082

7. Gulshan V, Peng L, Coram M, Stumpe MC, Wu D, Narayanaswamy A, Venugopalan S, Widner K, Madams T, Cuadros J, Kim R. Development and validation of a deep learning algorithm for detection of diabetic retinopathy in retinal fundus photographs. JAMA. 2016 Dec 13;316(22):2402–10.

8. Poplin R, Varadarajan AV, Blumer K, Liu Y, McConnell MV, Corrado GS, Peng L, Webster DR. Prediction of cardiovascular risk factors from retinal fundus photographs via deep learning. Nature Biomedical Engineering. 2018 Mar;2(3):158–64.

9. Campanella G, Hanna MG, Geneslaw L, Miraflor A, Werneck Krauss Silva V, Busam KJ, Brogi E, Reuter VE, Klimstra DS, Fuchs TJ. Clinical-grade computational pathology using weakly supervised deep learning on whole slide images. Nature Medicine. 2019 Aug;25(8):1301–9.

10. Jaroensri R, Wulczyn E, Hegde N, Brown T, Flament-Auvigne I, Tan F, Cai Y, Nagpal K, Rakha EA, Dabbs DJ, Olson N. Deep learning models for histologic grading of breast cancer and association with disease prognosis. NPJ Breast Cancer. 2022 Oct 4;8(1):1–2.

11. Tolkach Y, Dohmgörgen T, Toma M, Kristiansen G. High-accuracy prostate cancer pathology using deep learning. Nature Machine Intelligence. 2020 Jul;2(7):411–8.

12. Pierson E, Cutler DM, Leskovec J, Mullainathan S, Obermeyer Z. An algorithmic approach to reducing unexplained pain disparities in underserved populations. Nature Medicine. 2021 Jan;27(1):136–40.

13. Tiulpin A, Thevenot J, Rahtu E, Lehenkari P, Saarakkala S. Automatic knee osteoarthritis diagnosis from plain radiographs: a deep learning-based approach. Scientific reports. 2018 Jan 29;8(1):1–0.

14. Ueda D, Ehara S, Yamamoto A, Iwata S, Abo K, Walston SL, Matsumoto T, Shimazaki A, Yoshiyama M, Miki Y. Development and Validation of Artificial Intelligence–based Method for Diagnosis of Mitral Regurgitation from Chest Radiographs. Radiology: Artificial Intelligence. 2022 Mar 2;4(2):e210221.

15. Nam JG, Park S, Hwang EJ, Lee JH, Jin KN, Lim KY, Vu TH, Sohn JH, Hwang S, Goo JM, Park CM. Development and validation of deep learning–based automatic detection algorithm for malignant pulmonary nodules on chest radiographs. Radiology. 2019 Jan;290(1):218–28.

16. Courtiol P, Maussion C, Moarii M, Pronier E, Pilcer S, Sefta M, Manceron P, Toldo S, Zaslavskiy M, Le Stang N, Girard N. Deep learning-based classification of mesothelioma improves prediction of patient outcome. Nature medicine. 2019 Oct;25(10):1519–25.

17. Irvin J, Rajpurkar P, Ko M, Yu Y, Ciurea-Ilcus S, Chute C, Marklund H, Haghgoo B, Ball R, Shpanskaya K, Seekins J. Chexpert: A large chest radiograph dataset with uncertainty labels and expert comparison. In Proceedings of the AAAI Conference on Artificial Intelligence. 2019 Jul 17 (Vol. 33, No. 01, pp. 590–597).

18. Jaiswal AK, Tiwari P, Kumar S, Gupta D, Khanna A, Rodrigues JJ. Identifying pneumonia in chest X-rays: A deep learning approach. Measurement. 2019 Oct 1;145:511–8.

19. Chowdhury ME, Rahman T, Khandakar A, Mazhar R, Kadir MA, Mahbub ZB, Islam KR, Khan MS, Iqbal A, Al Emadi N, Reaz MB. Can AI help in screening viral and COVID-19 pneumonia?. IEEE Access. 2020 Jul 20;8:132665–76.

20. Lee JH, Sun HY, Park S, Kim H, Hwang EJ, Goo JM, Park CM. Performance of a deep learning algorithm compared with radiologic interpretation for lung cancer detection on chest radiographs in a health screening population. Radiology. 2020 Dec;297(3):687–96.

21. Yoo H, Kim KH, Singh R, Digumarthy SR, Kalra MK. Validation of a deep learning algorithm for the detection of malignant pulmonary nodules in chest radiographs. JAMA network open. 2020 Sep 1;3(9):e2017135-.

22. Hwang EJ, Park S, Jin KN, Im Kim J, Choi SY, Lee JH, Goo JM, Aum J, Yim JJ, Cohen JG, Ferretti GR. Development and validation of a deep learning–based automated detection algorithm for major thoracic diseases on chest radiographs. JAMA Network Open. 2019 Mar 1;2(3):e191095-.

23. Nabulsi Z, Sellergren A, Jamshy S, Lau C, Santos E, Kiraly AP, Ye W, Yang J, Pilgrim R, Kazemzadeh S, Yu J. Deep learning for distinguishing normal versus abnormal chest radiographs and generalization to two unseen diseases tuberculosis and COVID-19. Scientific Reports. 2021 Sep 1;11(1):1–5.

24. Tang YX, Tang YB, Peng Y, Yan K, Bagheri M, Redd BA, Brandon CJ, Lu Z, Han M, Xiao J, Summers RM. Automated abnormality classification of chest radiographs using deep convolutional neural networks. NPJ digital medicine. 2020 May 14;3(1):70.

25. Tavaziva G, Majidulla A, Nazish A, Saeed S, Benedetti A, Khan AJ, Khan FA. Diagnostic accuracy of a commercially available, deep learning-based chest X-ray interpretation software for detecting culture-confirmed pulmonary tuberculosis. International Journal of Infectious Diseases. 2022 Sep 1;122:15–20.

26. Qin ZZ, Ahmed S, Sarker MS, Paul K, Adel AS, Naheyan T, Barrett R, Banu S, Creswell J. Tuberculosis detection from chest x-rays for triaging in a high tuberculosis-burden setting: an evaluation of five artificial intelligence algorithms. The Lancet Digital Health. 2021 Sep 1;3(9):e543–54.

27. Majkowska A, Mittal S, Steiner DF, Reicher JJ, McKinney SM, Duggan GE, Eswaran K, Cameron Chen PH, Liu Y, Kalidindi SR, Ding A. Chest radiograph interpretation with deep learning models: assessment with radiologist-adjudicated reference standards and population-adjusted evaluation. Radiology. 2020 Feb;294(2):421–31.

28. Putha P, Tadepalli M, Reddy B, Raj T, Chiramal JA, Govil S, Sinha N, KS M, Reddivari S, Jagirdar A, Rao P. Can artificial intelligence reliably report chest x-rays?: Radiologist validation of an algorithm trained on 2.3 million x-rays. arXiv preprint arXiv:1807.07455. 2018 Jul 19.

29. Kazemzadeh S, Yu J, Jamshy S, Pilgrim R, Nabulsi Z, Chen C, Beladia N, Lau C, McKinney SM, Hughes T, Kiraly AP. Deep Learning Detection of Active Pulmonary Tuberculosis at Chest Radiography Matched the Clinical Performance of Radiologists. Radiology. 2022 Sep 6:212213.

30. Khan FA, Majidulla A, Tavaziva G, Nazish A, Abidi SK, Benedetti A, Menzies D, Johnston JC, Khan AJ, Saeed S. Chest x-ray analysis with deep learning-based software as a triage test for pulmonary tuberculosis: a prospective study of diagnostic accuracy for culture-confirmed disease. The Lancet Digital Health. 2020 Nov 1;2(11):e573–81.

31. US Department of State. Agency Information Collection Activities; Proposals, Submissions, and Approvals: Electronic Medical Examination for Visa or Refugee Applicant. https://www.regulations.gov/document/DOS_FRDOC_0001-5900 Date: 2022 Date accessed: June 30, 2023

32. OMB. DS-3030 Tuberculosis Worksheet: Medical Examination for Immigrant or Refugee Applicant. https://omb.report/icr/202010-1405-004/doc/105591400https://omb.report/icr/202010-1405-004/doc/105591400 Date: 2020. Date accessed: July 3, 2023

33. Wang X, Peng Y, Lu L, Lu Z, Bagheri M, Summers RM. Chestx-ray8: Hospital-scale chest x-ray database and benchmarks on weakly-supervised classification and localization of common thorax diseases. In Proceedings of the IEEE Conference on Computer Vision and Pattern Recognition. 2017 (pp. 2097–2106).

34. Jaeger S, Candemir S, Antani S, Wáng YX, Lu PX, Thoma G. Two public chest X-ray datasets for computer-aided screening of pulmonary diseases. Quantitative Imaging in Medicine and Surgery. 2014 Dec;4(6):475.

35. Nguyen, H.Q., Lam, K., Le, L.T. et al. VinDr-CXR: An open dataset of chest X-rays with radiologist’s annotations. Sci Data 9, 429 (2022). 10.1038/s41597-022-01498-w.

36. Tan M, Le Q. EfficientnetV2: Smaller models and faster training. In International Conference on Machine Learning. 2021 Jul 1 (pp. 10096–10106). PMLR.

37. Deng J, Dong W, Socher R, Li LJ, Li K, Fei-Fei L. Imagenet: A large-scale hierarchical image database. In 2009 IEEE conference on computer vision and pattern recognition 2009 Jun 20 (pp. 248–255). IEEE.

38. Kingma DP, Ba J. Adam: A method for stochastic optimization. arXiv preprint arXiv:1412.6980. 2014 Dec 22.

39. Efron B. Better bootstrap confidence intervals. Journal of the American Statistical Association. 1987 Mar 1;82(397):171–85.

40. Selvaraju RR, Cogswell M, Das A, Vedantam R, Parikh D, Batra D. Grad-cam: Visual explanations from deep networks via gradient-based localization. In Proceedings of the IEEE international conference on computer vision 2017 (pp. 618–626).

41. Kapishnikov A, Bolukbasi T, Viégas F, Terry M. Xrai: Better attributions through regions. In Proceedings of the IEEE/CVF International Conference on Computer Vision 2019 (pp. 4948–4957).

42. Codlin AJ, Dao TP, Vo LN, Forse RJ, Van Truong V, Dang HM, Nguyen LH, Nguyen HB, Nguyen NV, Sidney-Annerstedt K, Squire B. Independent evaluation of 12 artificial intelligence solutions for the detection of tuberculosis. Scientific reports. 2021 Dec 13;11(1):23895.

43. Kik SV, Gelaw SM, Ruhwald M, Song R, Khan FA, van Hest R, Chihota V, Nhung NV, Esmail A, Celina Garfin AM, Marks GB. Diagnostic accuracy of chest X-ray interpretation for tuberculosis by three artificial intelligence-based software in a screening use-case: an individual patient meta-analysis of global data. medRxiv. 2022 Jan 27:2022–01.

44. Arun N, Gaw N, Singh P, Chang K, Aggarwal M, Chen B, Hoebel K, Gupta S, Patel J, Gidwani M, Adebayo J. Assessing the trustworthiness of saliency maps for localizing abnormalities in medical imaging. Radiology: Artificial Intelligence. 2021 Oct 6;3(6):e200267.

45. Saporta A, Gui X, Agrawal A, Pareek A, Truong SQ, Nguyen CD, Ngo VD, Seekins J, Blankenberg FG, Ng AY, Lungren MP. Benchmarking saliency methods for chest X-ray interpretation. Nature Machine Intelligence. 2022 Oct;4(10):867–78.

46. Kagujje M, Kerkhoff AD, Nteeni M, Dunn I, Mateyo K, Muyoyeta M. The performance of computer-aided detection digital chest X-ray reading technologies for triage of active tuberculosis among persons with a history of previous tuberculosis. Clinical Infectious Diseases. 2023 Feb 1;76(3):e894–901.

## Supplemental References

Efron B. Better bootstrap confidence intervals. Journal of the American Statistical Association. 1987 Mar 1;82(397):171–85.

Majkowska A, Mittal S, Steiner DF, Reicher JJ, McKinney SM, Duggan GE, Eswaran K, Cameron Chen PH, Liu Y, Kalidindi SR, Ding A. Chest radiograph interpretation with deep learning models: assessment with radiologist-adjudicated reference standards and population-adjusted evaluation. Radiology. 2020 Feb;294(2):421–31.

